# Sensitive one-step isothermal detection of pathogen-derived RNAs

**DOI:** 10.1101/2020.03.05.20031971

**Authors:** Chang Ha Woo, Sungho Jang, Giyoung Shin, Gyoo Yeol Jung, Jeong Wook Lee

## Abstract

The recent outbreaks of Ebola, Zika, MERS, and SARS-CoV-2 (2019-nCoV) require fast, simple, and sensitive onsite nucleic acid diagnostics that can be developed rapidly to prevent the spread of diseases. We have developed a SENsitive Splint-based one-step isothermal RNA detection (SENSR) method for rapid and straightforward onsite detection of pathogen RNAs with high sensitivity and specificity. SENSR consists of two simple enzymatic reactions: a ligation reaction by SplintR ligase and subsequent transcription by T7 RNA polymerase. The resulting transcript forms an RNA aptamer that induces fluorescence. Here, we demonstrate that SENSR is an effective and highly sensitive method for the detection of the current epidemic pathogen, *severe acute respiratory syndrome-related coronavirus* 2 (SARS-CoV-2). We also show that the platform can be extended to the detection of five other pathogens. Overall, SENSR is a molecular diagnostic method that can be developed rapidly for onsite uses requiring high sensitivity, specificity, and short assaying times.

## Introduction

Increasing global trade and travel are considered the cause of frequent emergence and rapid dissemination of infectious diseases around the world. Some life-threatening infectious diseases often have signs and symptoms similar to cold or flu-like syndromes. Early diagnosis is therefore essential to identify the diseases and provide the correct treatment. Immediate and onsite diagnostic decisions also help to prevent the spread of epidemic and pandemic infectious diseases^1–3^. In order to rapidly diagnose infectious diseases, a nucleic acid-based diagnosis has emerged as an alternative to the conventional culture-based, or immunoassay-based, approaches due to their rapidity or specificity^4–6^.

To increase sensitivity, current nucleic acid detection methods generally involve a target amplification step prior to the detection step. The conventional amplification method is based on PCR, which requires a thermocycler for delicate temperature modulation. As an alternative to the thermal cycling-based amplification, isothermal amplification methods are available, which rely primarily on a strand-displacing polymerase or T7 RNA polymerase at a constant temperature^7^. However, the complex composition of the isothermal amplification mixtures often renders these approaches incompatible with detection methods and whole diagnosis generally becomes a multi-step process^8–11^. The diagnostic regimen with multi-step procedures requires additional time, instruments, and reagents, as well as skilled personnel to perform the diagnostic procedure. This aspect limits the broad applicability of nucleic acid diagnostics, especially in situations where rapid and simple detection is required.

Ligation-dependent nucleic acid detection is a sequence-specific method that primarily depends on ligation of two separate probes that hybridize to adjacent sites of the target sequence^12^. Because of their specificity, the ligation-dependent methods are used for detection of markers of genetic disorders^13,14^ and pathogens^15,16^, typically combined with subsequent amplification and signal generation methods. In particular, the SplintR ligase can efficiently ligate two DNA probes using a target single-stranded RNA as a splint, enabling the sequence-specific detection of RNA molecule^17,18^. Because the reaction components of the ligation-dependent methods are relatively simple, we hypothesized that the ligation-dependent method could be exploited to establish a one-step RNA detection platform when combined with compatible amplification and signal generation methods in a single reaction mixture.

In this study, we developed a one-step, ligation-dependent isothermal reaction cascade that enables rapid detection of RNAs with high sensitivity, termed SENsitive Splint-based one-step isothermal RNA detection (SENSR). SENSR consists of two simple enzymatic reactions, a ligation reaction by SplintR ligase and subsequent transcription by T7 RNA polymerase. The resulting transcript forms an RNA aptamer that binds to a fluorogenic dye and produces fluorescence only when target RNA exists in a sample. SENSR was able to detect target RNA of Methicillin-Resistant *Staphylococcus aureus* (MRSA) in 30 minutes with a limit of detection of 0.1 aM. We further applied this platform to detect various pathogens, *Vibrio vulnificus, Escherichia coli* O157:H7, Middle East Respiratory Syndrome-related Coronavirus (MERS-CoV), and Influenza A viruses, by merely redesigning the hybridization regions of the probes. Finally, we demonstrated the fast development of the SENSR assay for the latest epidemic pathogen, *Severe acute respiratory syndrome-related coronavirus* 2 (SARS-CoV-2 or 2019-nCoV), using minimal, publicly available information.

## Results

### Design of one-step, isothermal reaction cascade

We designed a reaction cascade that allows the one-step diagnostic test, in which all reaction steps for nucleic acid detection occur simultaneously in a single tube (Fig. 1). The cascade consists of four core components, which includes only two enzymes: a set of oligonucleotide probes, SplintR ligase, T7 RNA polymerase, and a fluorogenic dye. The components were mixed in a buffer solution with ribonucleotides. Upon addition of the pathogen-derived RNA sample, the reaction steps of ligation, transcription, and dye-aptamer binding enabled detection, amplification, and signal production, respectively.

**Fig. 1:**
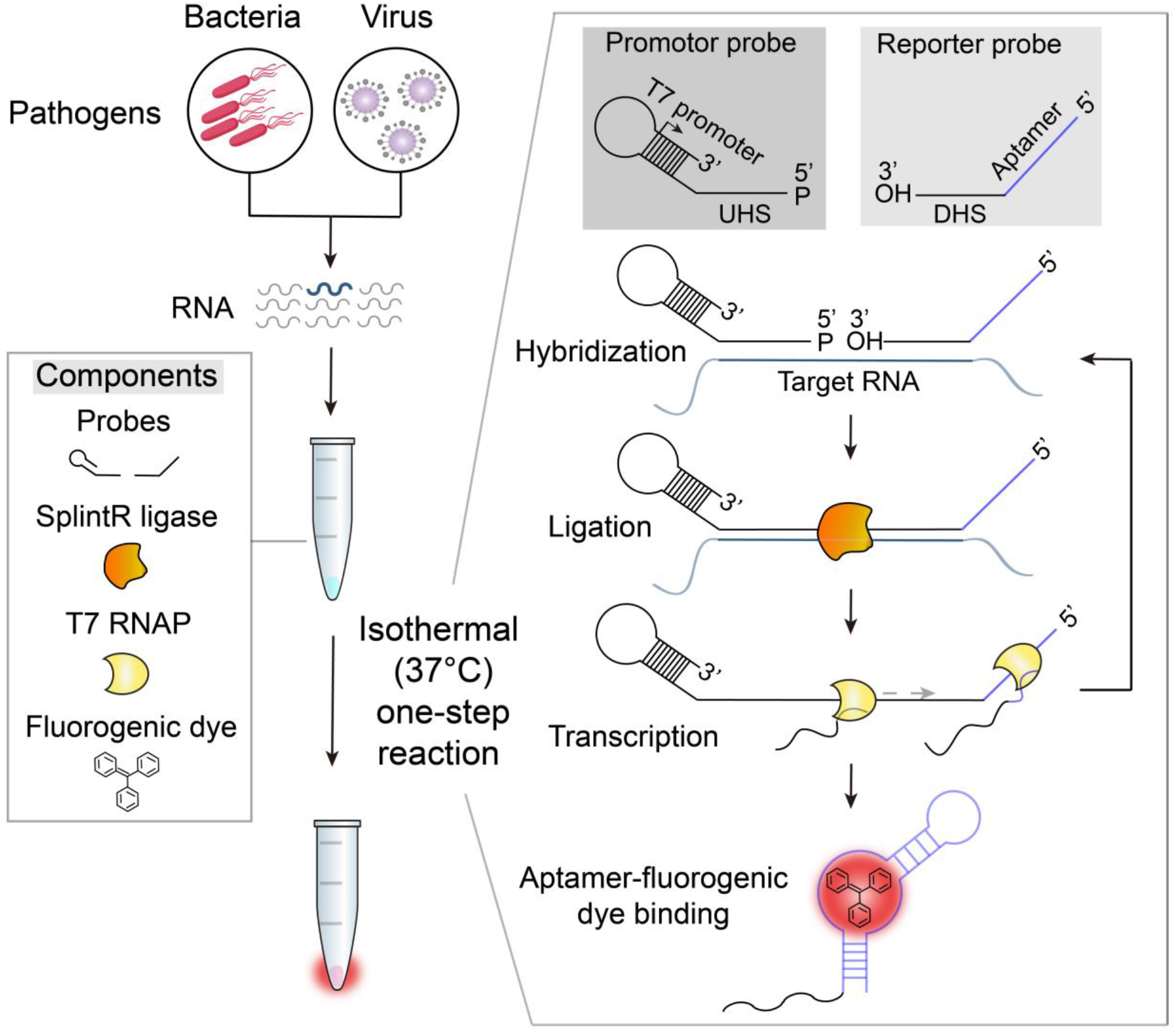
Schematic illustration of SENSR, a one-step isothermal reaction cascade for rapid detection of RNAs. The reaction is composed of four main components: a set of probes, SplintR ligase, T7 RNA polymerase (T7 RNAP), and a fluorogenic dye. In the presence of target RNA, hybridization, ligation, transcription, and aptamer-dye binding reactions occur sequentially in a single reaction tube at a constant temperature. UHS, upstream hybridization sequence; DHS, downstream hybridization sequence.

Two single-stranded DNA probes were designed to include several functional parts involved in amplification, detection, and signal generation, thereby eliminating the need for human intervention during the entire diagnostic process (Fig. 1). First, the promoter probe consists of an upstream hybridization sequence (UHS) and a stem-loop T7 promoter. The UHS hybridizes to the 5′-half of a target RNA region. The stem-loop T7 was adopted from the literature^19^ (Supplementary Table 1) to form an active, double-stranded T7 promoter using a single-stranded oligonucleotide. The sequence of the UHS was designed by Primer-BLAST^20^ to ensure specific binding to the target RNA. Among candidate UHS sequences, we chose the one with minimal secondary structure at 37 °C predicted by NUPACK^21^ to maximize the hybridization between the UHS and its target region (Supplementary Table 2). The 5′-end of the promoter probe was then phosphorylated for ligation. Next, a reporter probe consists of a downstream hybridization sequence (DHS) and a template sequence for a dye-binding RNA aptamer. The DHS contains the complementary sequence to the other half of the target RNA region. Similar to the UHS, the DHS was selected to have minimal predicted secondary structure (Supplementary Table 2).

Once both UHS and DHS probes hybridize correctly to the target RNA, SplintR ligase can initiate the cascade by connecting the probes that have all features built for the one-step diagnostic test. Subsequently, T7 RNA polymerase can synthesize the RNA aptamer using the full-length, ligated probe as a DNA template, which can be bound with the fluorogenic dye to emit fluorescence as an output (Fig. 1). Notably, the reaction scheme of SENSR inherently supports two mechanisms that could amplify the signal: 1) multiple transcription events from the full-length, ligated probe by T7 RNA polymerase and 2) the presence of target RNA sequence on the full-length transcript which could be utilized as an additional splint for unligated probes in the reaction mixture. Accordingly, SENSR could enable sensitive RNA detection without any pre-amplification steps.

### Construction of each component reaction in SENSR

In this study, we used MRSA as a model case to validate each reaction step that constitutes SENSR. MRSA is of particular interest because it requires significant effort to minimize healthcare-related infections and prevent future infectious diseases of drug-resistant pathogens^22^.

First, we designed a pair of probes that target the *mecA* transcript of MRSA following the probe design process described in the previous section (Supplementary Note 1 and Supplementary Tables 1 and 3), and the RNA-splinted ligation between the two probes was tested. The probes were ligated using SplintR ligase with or without the target RNA, and the reaction resultants were further amplified with a pair of PCR primers and analyzed (see Methods section). The correct size of the PCR product was obtained only when the two probes and target RNA were added together to the ligation mixture (Fig. 2a). This result indicates that our probes were successfully ligated only in the presence of the target RNA.

**Fig. 2:**
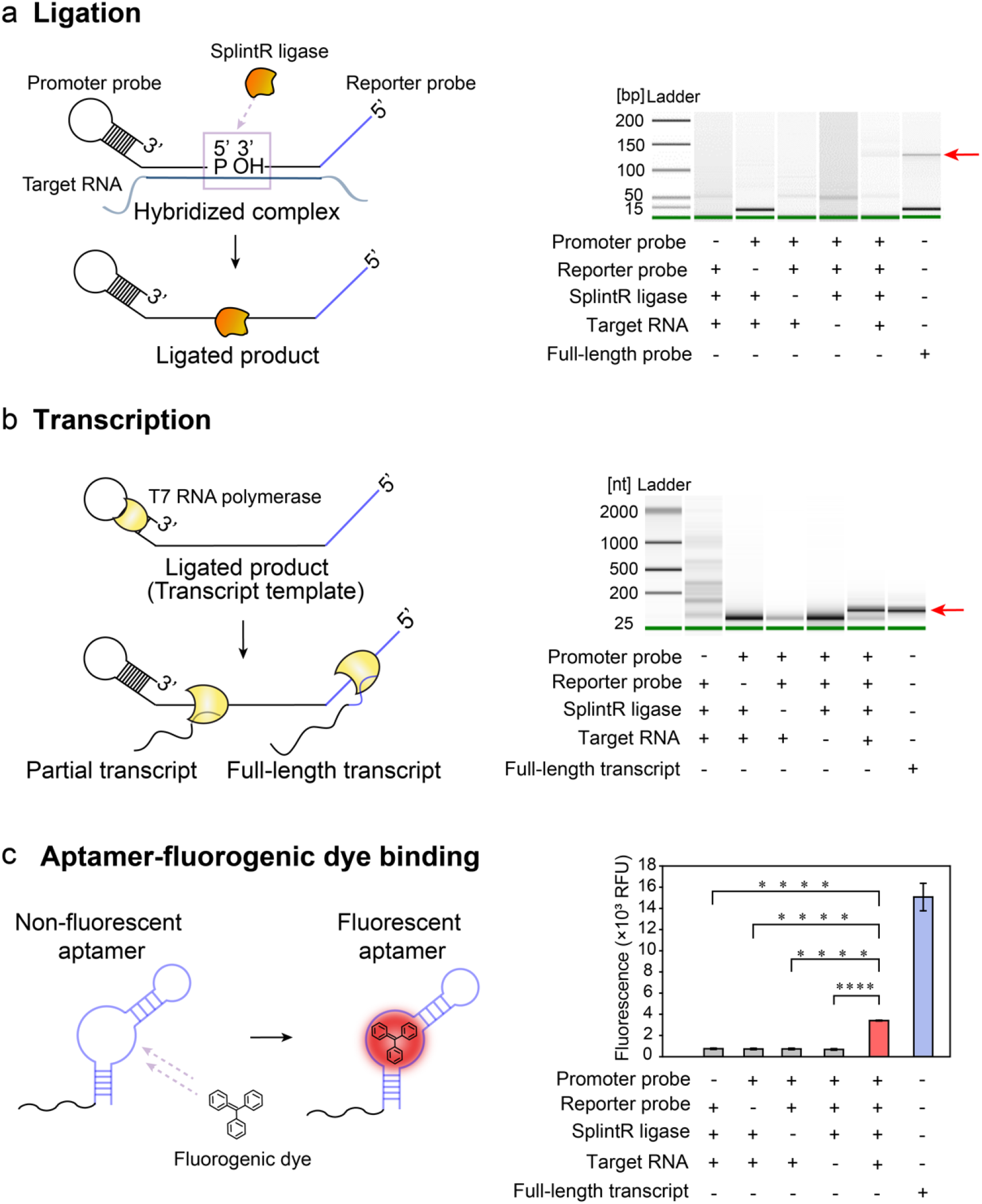
Construction of the three components reactions of SENSR. **a**, Ligation reaction. The ligation resultants were amplified with a pair of PCR primers (LigChk_F,R in Supplementary Table 4) and analyzed using Bioanalyzer. The ligation reaction occurred when only the promoter probe, reporter probe, SplintR ligase, and target RNA were all present. A full-length probe combining the promoter and reporter probes was amplified with the same set of PCR primers and used as a size control, as indicated by the red arrow. **b**, Transcription reaction. The ligated mixtures were used as a DNA template to validate transcription. The transcript was obtained only in the presence of target RNA and all other components, demonstrating both target-dependent ligation and the subsequent transcription. The red arrow points to the correct size of the transcript. **c**, Fluorescence reaction. After sequential ligation and transcription reactions, the reaction mixture with the correct size of the transcript produced higher fluorescence compared to other conditions that lack one of the necessary components. Fluorescence tests are four experimental replicas (two-tailed student’s test; * P < 0.05, ** P < 0.01, *** P < 0.001, **** P < 0.0001; bars represent mean ± s.d).

We then used the ligated probe as a DNA template to test whether transcription could occur. The ligation mixture was added at a 1/10 ratio to the *in vitro* transcription reaction mixture with T7 RNA polymerase. Only when the target RNA was present in the ligation reaction was the full-length transcript (92 nt) observed from transcription, thereby confirming both target-dependent ligation and the subsequent transcription (Fig. 2b).

Finally, we confirmed that the transcript from the full-length ligated probe could produce fluorescence upon binding to the fluorogenic dye. The reaction mixture of sequential ligation and transcription reactions were purified, and an equal amount of RNAs from each combination was incubated with the fluorogenic dye (Fig. 2c). The RNA product from the reaction mixture with the two probes and target RNA produced higher fluorescence than that of the other combinations. Therefore, we confirmed that the target RNA could be detected using a set of probes by performing each component reaction in SENSR.

### Development of one-step isothermal reaction cascade

Since all component reactions in SENSR were validated in their respective buffers, we then sought to develop a one-step reaction condition with a single reaction buffer at a single temperature, where all reaction steps, including probe annealing, ligation, transcription, and aptamer fluorescence reaction occur simultaneously. To accomplish this, we first investigated a wide range of temperatures (25–95 °C) for hybridization of the probes and target. Then, each mixture was subjected to the sequential ligation, transcription reactions, and fluorescence reaction described in the previous section. Remarkably, fluorescence was observed at all hybridization temperatures, including 35 °C and 40 °C (Supplementary Fig. 1), the optimal temperature ranges for enzyme activities in SENSR, thereby suggesting that the entire reaction can be built up as an isothermal reaction.

Additionally, a single reaction buffer composition suitable for all reaction steps was configured to establish all reactions in one pot. Since T7 RNA polymerase reaction buffer has the most inclusive composition of the four reaction buffers (probe annealing, ligation, transcription, and aptamer fluorescence reaction buffers) we used T7 RNA polymerase buffer as a basis for the optimization. Various reaction conditions were optimized, including the reaction temperature and concentrations of various enzymes, and components to enhance the fluorescence signal (Supplementary Note 2 and Supplementary Figs. 2 and 3). The optimized SENSR condition enabled the detection of target RNA in a one-pot isothermal reaction at 37 °C.

**Fig. 3:**
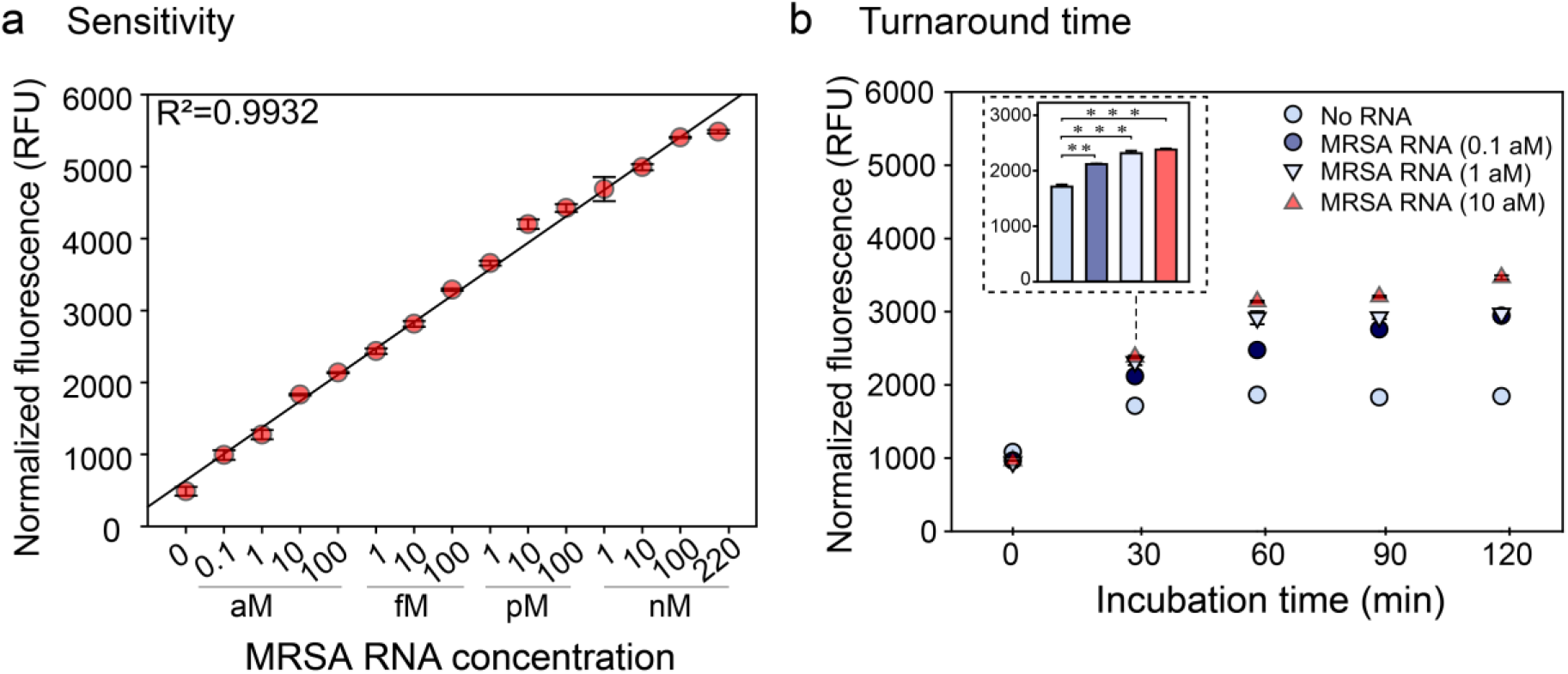
Sensitivity and turnaround time of SENSR. **a**, Sensitivity of SENSR. The target RNA from 220 nM to 0.1 aM was tested. The detection limit is 0.1 aM. High linearity suggests that SENSR can be used for the quantification of the target RNA. **b**, Turnaround time of SENSR. To check the time required for the SENSR reaction, the incubation time of SENSR was varied. The target RNA of 0.1 aM was detected as early as 30 min. All tests are four experimental replicas (two-tailed student’s test; * P < 0.05, ** P < 0.01, *** P < 0.001, **** P < 0.0001; bars represent mean ± s.d).

### Rapid and sensitive RNA detection by SENSR

Since the one-step and one-pot isothermal reaction condition was established, we then assessed the sensitivity and turnaround time of SENSR. We evaluated the sensitivity by measuring fluorescence from one-step reactions containing the *mecA* probe pair and synthetic *mecA* RNA in the range of 0.1 aM to 220 nM (Fig. 3a). Notably, the detection limit was as low as 0.1 aM (corresponding to 6 molecules per 100 µ L reaction), indicating the high sensitivity of SENSR. Moreover, the linearity of the fluorescence intensity over a wide range of concentrations (R^2^ = 0.9932) suggests that SENSR can be used for target RNA quantification.

We then measured the minimal turnaround time required to confirm the presence of the target RNA in a sample. The target RNA ranging from 0.1 aM to 10 aM were added to the SENSR reaction, and fluorescence was measured every 30 minutes. The fluorescence with 0.1 aM was discernible against the negative control in only 30 minutes (Fig. 3b). Further incubation of the reaction better distinguished the target RNA-containing reaction from the negative control reaction. Collectively, the SENSR reaction was able to specifically detect the target RNA within 30 minutes with a detection limit of 0.1 aM.

### Broad adaptability of SENSR for pathogen detection

With the fast and sensitive RNA detection using SENSR, we next attempted to reconfigure this platform for the detection of RNA markers from various pathogens. Target RNA sequences for SENSR are specified by only two hybridization regions (UHS and DHS) of probes, which makes the probe design process fast and straightforward without many computational steps. This design feature, requiring only nucleotide sequences to build molecular diagnostics, allows for easy construction of SENSR probes for any infectious diseases (Fig. 4a).

**Fig. 4:**
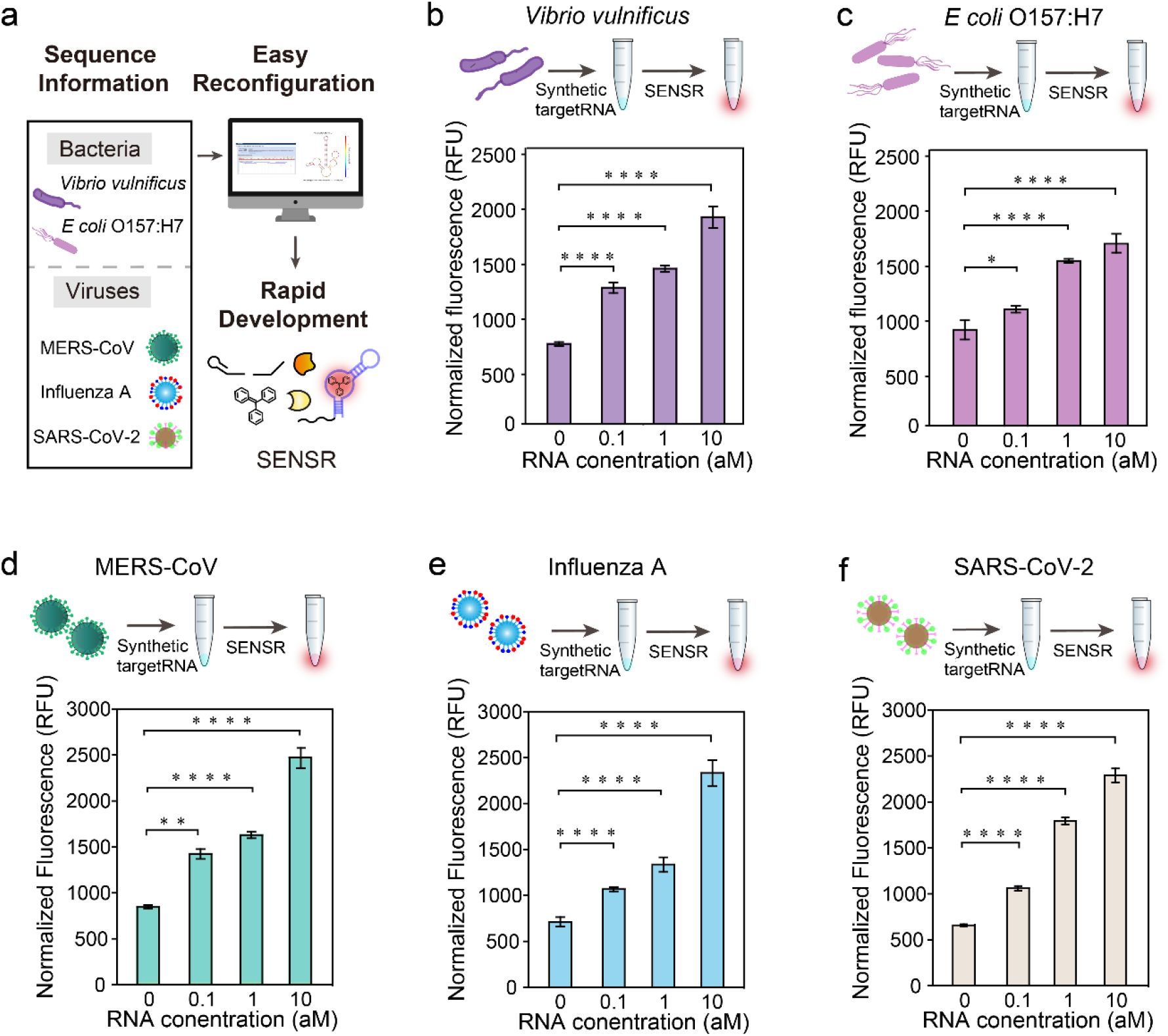
Broad adaptability of SENSR. Two pathogenic microbes and three viruses were targeted by redesigning probe sequences. **a**, Schematic of SENSR with easy reconfiguration and rapid development. **b**,**c**, Detection of bacterial RNA markers, for *V. vulnificus* and *E. coli O157:H7*, respectively. **d**,**e**,**f**, Detection of viral RNA markers, MERS-CoV, Influenza A, and SARS-CoV-2, respectively. All probe pairs tested showed high sensitivity and linearity to detect RNA markers. All tests are four experimental replicas (two-tailed student’s test; * P < 0.05, ** P < 0.01, *** P < 0.001, **** P < 0.0001; bars represent mean ± s.d).

To demonstrate SENSR for various pathogens, we first targeted two pathogenic microorganisms, *V. vulnificus* and *E. coli* O157:H7. *V. vulnificus* is known to cause gastroenteritis, wound infection, and sepsis in humans. We designed a probe pair targeting *vvhA* (Supplementary Table 3), a *V. vulnificus*-specific target encoding extracellular hemolysin with hemolytic activity and cytotoxic effect. The sensitivity of SENSR reaction using the probe pair and *in-vitro*-transcribed *vvhA* RNA was as low as 0.1 aM (Fig. 4b), and a linear correlation was observed between the concentration of RNA and the fluorescence intensity (R^2^=0.9566).

Next, we used SENSR to detect *E. coli* O157:H7, which causes foodborne illness. Similar to that for *V. vulnificus*, we designed a probe pair for *E. coli* O157:H7-specific target gene, *tir* (Supplementary Table 3), for SENSR reaction. Similarly, RNA concentrations as low as 0.1 aM were detected by SENSR and a high linear correlation between the concentration of RNA and the fluorescence intensity was observed (Fig. 4c; R^2^=0.9684).

The target was expanded to human-infective RNA viruses that cause fatal diseases^23^. First, we aimed at Middle East Respiratory Syndrome-related Coronavirus (MERS-CoV). The mortality rate of MERS was reported to be 35%^24^, and can be transmitted from human to human^25^, which raises the need for a fast and sensitive onsite diagnostic test. The probe pairs for the MERS-specific gene, *upE* (Supplementary Table 3), exhibited similar sensitivity and linearity to the bacterial cases (Fig. 4d). Likewise, we designed a probe pair for the Influenza A virus-specific target gene, HA (hemagglutinin) gene (Supplementary Table 3). SENSR was able to detect the Influenza A RNA target with similar sensitivity and linearity (Fig. 4e). Finally, we designed a probe pair for a recently emerging pathogen, SARS-CoV-2. The target sequence was selected based on the standard real-time RT-PCR assay for the SARS-CoV-2^26^, which aimed at the RNA-dependent RNA polymerase (*RdRp*) gene (Supplementary Table 3). Again, SENSR successfully detected its target RNA as low as 0.1 aM, which corroborates the high adaptability of this method to various RNA markers (Fig. 2f).

Taken together, we demonstrated that SENSR could be easily reconfigured to detect various RNA markers of pathogens by redesigning the probes. The probe design process is simple and requires a small amount of computation using open web-based software. All probe pairs tested showed high sensitivity and linearity for detecting RNA markers, reinforcing the robustness of the probe design process and the wide expandability of SENSR.

### Direct detection of a pathogen using SENSR

Next, we employed SENSR for the detection of RNA samples derived from the live cells of a pathogen. We targeted MRSA, whose marker RNA was detected by SENSR. Methicillin-Sensitive *Staphylococcus aureus* (MSSA) that contains no target mRNA was used as a negative control. MRSA and MSSA cells were heated to 95 °C to lyse the cells and to release RNAs. The samples were then diluted and added to SENSR reaction to investigate the specificity and sensitivity (Fig. 5a). We observed a significant difference in fluorescence intensity between MRSA and MSSA (Fig. 5b). The RNA sample from only 2 CFU per 100 μL reaction of MRSA, not MSSA, was clearly detected by SENSR, thereby indicating its high sensitivity and specificity even with samples of the living pathogen. Finally, the performance of SENSR was further validated using samples prepared in human serum (Fig. 5c). The sensitivity and specificity of SENSR were unaffected by the presence of human serum (Fig. 5d), indicating the suitability of SENSR in practical applications.

**Fig. 5:**
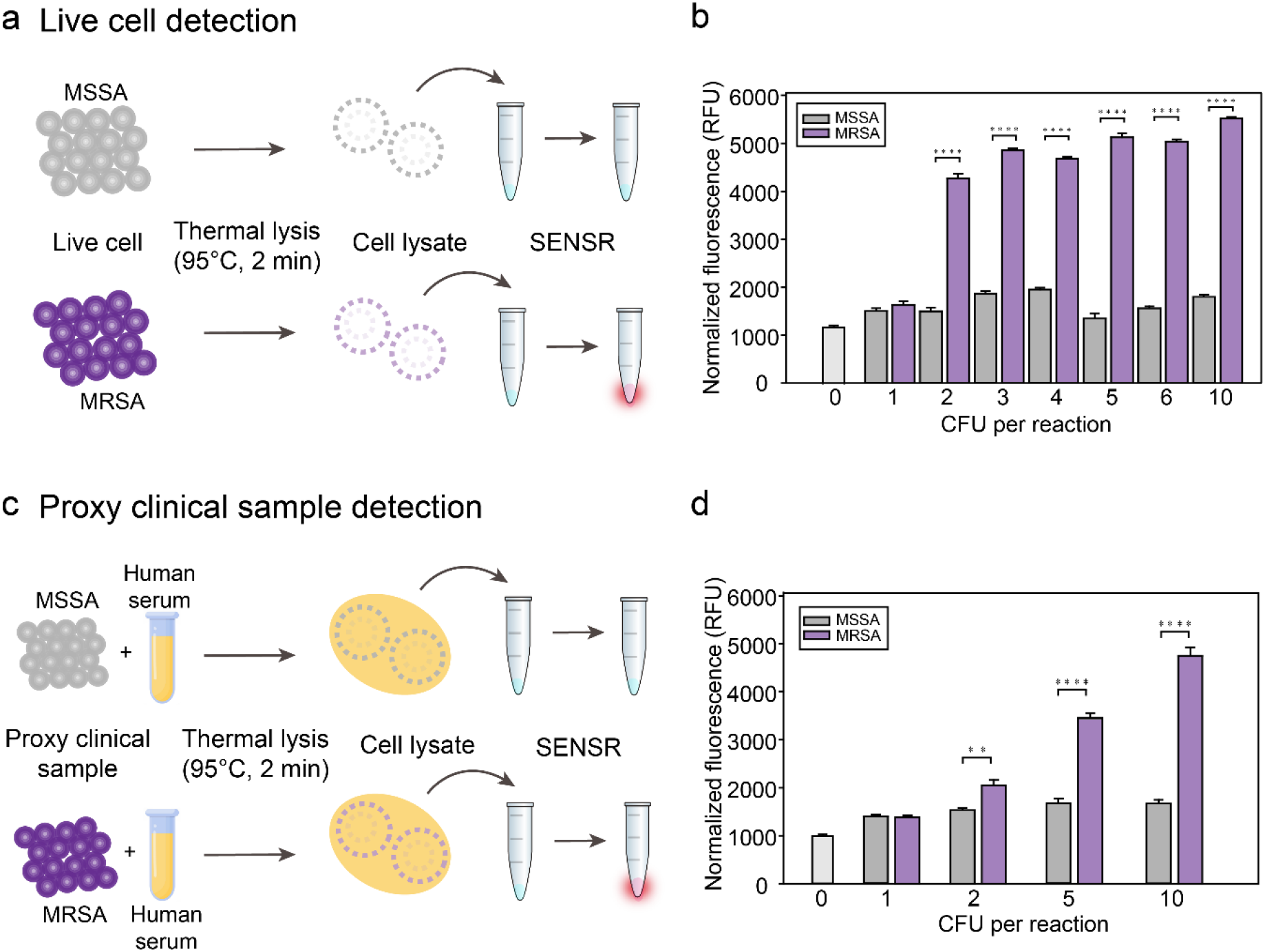
Live cell and proxy clinical sample detection using SENSR. **a**, Direct detection of bacterial cells. Thermal cell lysates of MRSA and MSSA were subjected to the SENSR reaction. **b**, Clear distinction in the fluorescence intensity between MRSA and MSSA samples. The detection limit of SENSR is as low as 2 CFU per 100 µ L reaction. **c**, Detection of bacterial cell diluted in human serum as a proxy clinical sample. Bacteria-contained human serum was thermally lysed and subjected to the SENSR reaction. **d**, An obvious distinction in the fluorescence intensity between MRSA- and MSSA-contained human serum was observed. The detection limit of SENSR is as low as 2 CFU per 100 µ L reaction. All tests are four experimental replicas (two-tailed student’s test; * P < 0.05, ** P < 0.01, *** P < 0.001, **** P < 0.0001; bars represent mean ± s.d).

### Dual target detection using orthogonal SENSR probes

Finally, we expanded the capability of SENSR to enable the simultaneous detection of two target RNAs in a single reaction by leveraging simple probe design. The detection of multiple biomarkers is frequently needed to make a better decision by reducing false-positive and false-negative results. Based on the high specificity of SENSR probes and availability of light-up RNA aptamers with distinct spectral properties^27^, we hypothesized that we could design two sets of SENSR probes that operate orthogonally in a single reaction to detect their respective target RNAs.

First, we developed an orthogonal reporter probe for Influenza A virus. Since the MRSA infection causes flu-like symptoms, discrimination of this pathogen from common Influenza A virus is required. Furthermore, the patient with the influenza A infection is more susceptible to the MRSA infection^28^. Collectively, simultaneous detection and discrimination of both pathogens can help the diagnosis and follow-up actions. An orthogonal reporter probe for Influenza A virus was designed by replacing its aptamer template region with the template for the Broccoli aptamer^29–32^ which binds to DFHBI-1T ((5Z)-5-[(3,5-Difluoro-4-hydroxyphenyl)methylene]-3,5-dihydro-2-methyl-3-(2,2,2-trifluoroethyl)-4H-imidazol-4-one) and exhibits spectral properties distinct from that of the malachite green aptamer. Secondary structures of the new reporter probe and corresponding full-length RNA transcript were simulated, using NUPACK, and satisfied the probe design criteria without further optimization (Supplementary Table 2). Dual detection of MRSA and Influenza A virus was tested in SENSR reactions in which the two probe pairs, their cognate fluorogenic dyes, and various concentrations of the target RNAs were added (Fig. 6a). When the probe pairs are hybridized to their respective target RNAs, and successful transcription follows, the RNA aptamers would bind their cognate dyes and emit distinguishable fluorescence. The presence of each target RNA could be determined by the fluorescence patterns from the SENSR reaction: malachite green aptamer fluorescence for MRSA, and Broccoli aptamer fluorescence for Influenza A virus. Indeed, the presence of either target RNA (1 nM) was easily detected by the fluorescence pattern (Fig. 6b). Across various concentrations of each target RNA, the SENSR probes specifically produced fluorescence that responded only to respective targets, thereby enabling orthogonal dual detection of two pathogens (Fig. 6c).

**Fig. 6:**
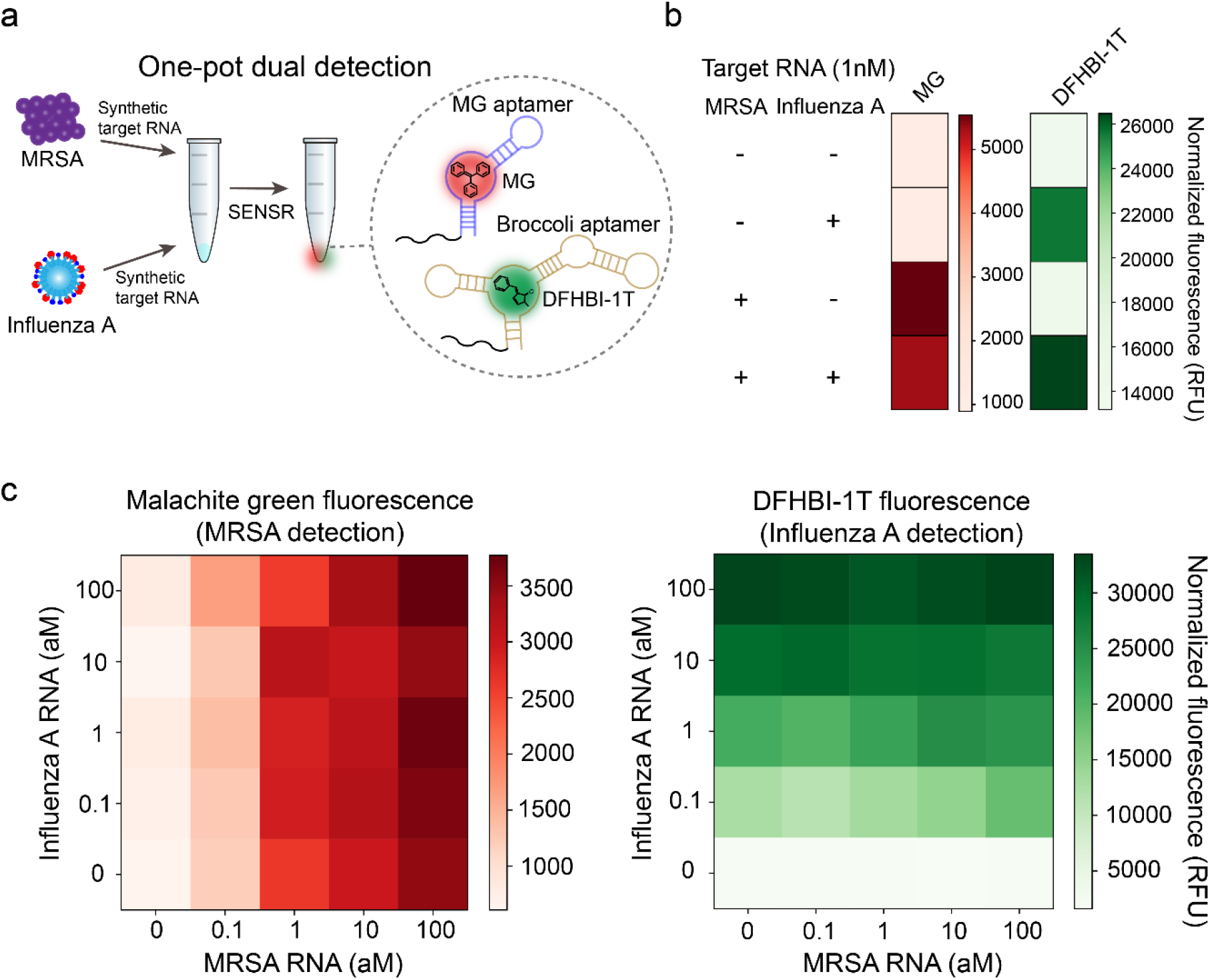
One-pot dual detection of RNAs by SENSR. **a**, One-pot dual detection of MRSA and Influenza A. The dual SENSR mixture contains two orthogonal pairs of probes and fluorogenic dyes with other components. Each probe pair hybridizes to the corresponding target RNA and allows SENSR reaction, emitting fluorescence that is distinguishable from each other. **b**, Validation of orthogonal dual SENSR reaction. Presence of each target RNA (1 nM) was determined by the intensities of non-overlapping fluorescence. **c**, One-pot dual SENSR detection of MRSA and Influenza A with various concentration combinations. All tests are four experimental replicas.

Lastly, we applied the orthogonal dual detection to the SARS-CoV-2, which has many related viruses with high sequence homology. Simultaneous detection of multiple target sites along its genome would enable specific discrimination of this emerging pathogen from others. In addition to the previously demonstrated SARS-CoV-2 probe pair (Fig. 4f), we designed three additional probe pairs for other regions in the *RdRp* gene with either the malachite green aptamer or the Broccoli aptamer (Fig. 7a). Each probe pair contained a discriminatory base at either the 5′-end of PP or 3′-end of RP, which are unique to SARS-CoV-2 against other similar viruses. Mismatches between the probes and nontarget RNAs would inhibit ligation and subsequent SENSR reaction and could enable more specific detection of SARS-CoV-2. Indeed, all four probe pairs were able to detect 1 aM of SARS-CoV-2 RNA, thereby exhibiting higher fluorescence intensity compared to that of the related viral RNA sequences (Fig. 7b). We then tested the orthogonal dual detection of two target regions using the SARS-CoV-2-MG1 and SARS-CoV-2-BR2 probe pairs. Dual SENSR assay effectively detected the target RNA and maintained the specificity of each probe pair (Fig. 7c). Therefore, the dual SENSR assay could be used to assist diagnostic decision making by providing two detection results that can complement each other.

**Fig. 7:**
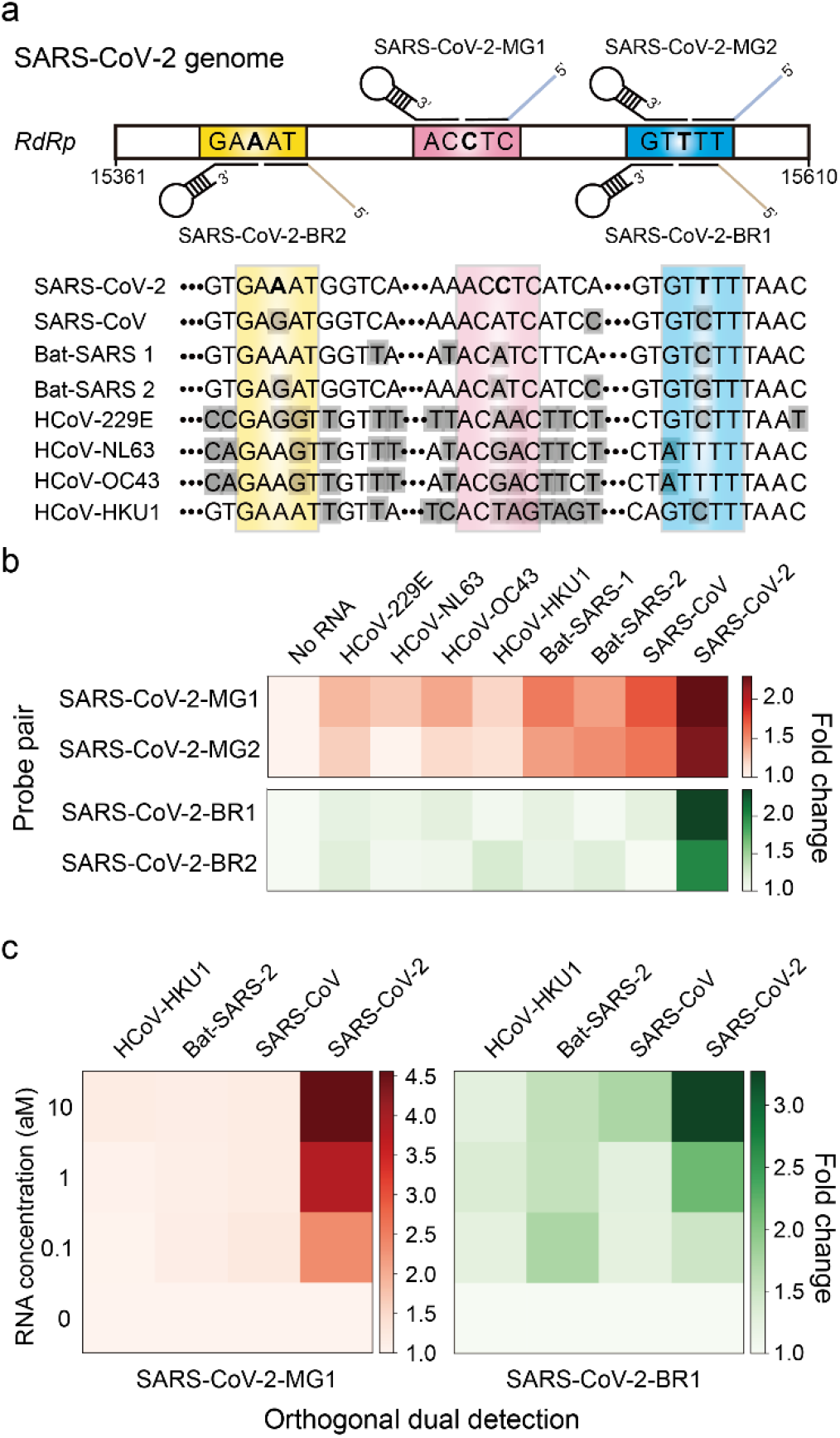
Dual detection of SARS-CoV-2 by SENSR. **a**, Probe design for dual SENSR detection. Three regions in the RNA-dependent RNA polymerase (*RdRp*) gene of SARS-CoV-2 were targeted. Discriminatory bases that enable specific detection of SARS-CoV-2 against viruses with highly similar sequences are marked by bold letters. Grey shades indicate mismatches between the sequences of SARS-CoV-2 and other viruses. **b**, Singleplex detection of 1 aM SARS-CoV-2 RNA by SENSR. 229E, *Human coronavirus* 229E; NL63, *Human coronavirus* NL63; OC43, *Human coronavirus* OC43; HKU1, *Human coronavirus* HKU1; Bat-SARS-1, Mg772933 *Bat SARS-related coronavirus*; Bat-SARS-2, NC_014470 *Bat SARS-related coronavirus*; SARS-CoV, *Severe acute respiratory syndrome-related coronavirus*; SARS-CoV-2, *Severe acute respiratory syndrome-related coronavirus* 2. **c**, One-pot dual detection of SARS-CoV-2 by orthogonal probe pairs, SARS-CoV-2-MG1 and - BR1. All tests are two experimental replicas. Fold changes were calculated by dividing the normalized fluorescence values by that with no target RNA.

## Discussion

Rapid, simple, economical, and sensitive diagnostic tests are needed to detect and manage infectious diseases at the earliest possible time. However, conventional approaches lack one or more of these features. Culture-based methods are time-consuming (>24 h)^33^ while PCR-based methods, including real-time-PCR, require a complex procedure, expensive instruments, and skilled expertise. Various isothermal amplification methods for RNA have been introduced to replace traditional methods^7,8^, but they generally require numerous reaction components, often making them expensive and incompatible with the signal production step.

In contrast, SENSR satisfies many desirable requirements for onsite diagnostic tests for pathogens, such as short turnaround time (30 min), low limit of detection (0.1 aM), inexpensive instrumentation and reagents, and a simple diagnostic procedure. SENSR integrates all component reaction steps using the specially designed probes that contain all required functional parts: promoter, hybridization sequence to target, and an aptamer template. Even with the multifaceted features of the SENSR probes, the design process is systematic and straightforward. Therefore, new SENSR assay can be promptly developed for emerging pathogens as exemplified by the successful design of SENSR assay for SARS-CoV-2.

The probe design is unique in that two DNA probes are designed to expose single-stranded target recognition parts, enabling hybridization of the target RNA and the probes at 37°C. The hybridization sequences were systematically selected using the nucleic acid design software Primer-BLAST and NUPACK to minimize any structure formation while maximizing hybridization to the target RNA. The efficient hybridization between the probes and target RNA is one of the reasons for enabling high sensitivity during the isothermal reaction.

The promoter probe is programmed to form a stem-loop structure and the stem part forms a double-stranded T7 promoter sequence that initiates transcription by recruiting T7 RNA polymerase. Since the two strands of T7 promoter part are physically connected by the loop, the probability of formation of a functional double-stranded promoter is higher in the stem-loop structured design than when each strand of the promoter is not connected by the loop. Thus, the hairpin structured, self-assembling promoter sequence in the promoter probe can facilitate hybridization and subsequent transcription more efficiently.

The initiated transcription elongates through the single-stranded DNA as a template to amplify target RNAs containing aptamer RNAs. The use of a fluorogenic RNA aptamer facilitated SENSR development by enabling fast and straightforward signal generation. Compared to conventional fluorescent protein outputs, the use of RNA aptamers as reporters can reduce the time it takes to observe the signal^34^.

The simple enzyme composition is another reason to enable one-step and one-pot detection. The fewer the enzymes, the easier it is to optimize in terms of temperature and buffer composition. In designing the detection scheme, we deliberately tried to reduce the number of enzymes, thus creating one of the simplest isothermal detection schemes based on two enzymes: SplintR ligase for target detection and T7 RNA polymerase for amplification.

In addition to the results shown in this study, we expect that SENSR has a broad range of possibilities for pathogen detection. First, SENSR can be easily implemented in the initial screening of infectious diseases at places where a large number of people gather and transfer^35,36^. With a short turnaround time and a simple reaction composition, SENSR is an ideal diagnostic test for rapid and economical screening. Second, SENSR will be a valuable platform for the immediate development of diagnostic tests for emerging pathogens^1,37^ because of the simple probe design process and broad adaptability of SENSR. In this work, we demonstrated the successful application of SENSR to six pathogens, using minimal redesign based on the highly modular structure of the probes. In theory, SENSR detection probes can be designed for any RNA as long as the target nucleic acid sequence is available. This feature provides SENSR a significant advantage over antibody-based diagnostics to rapidly respond to the outbreak of infectious disease. The nucleic acid probe synthesis is more scalable than animal antibody production. Therefore, SENSR is more suitable for rapid mass production of diagnostic kits than antibody-based diagnostics. Future efforts on automated probe design will be needed to accelerate the development of SENSR assays for newly emerging pathogens.

In conclusion, SENSR is a powerful diagnostic platform for RNA detection, which offers a short turnaround time, high sensitivity and specificity, and a simple assay procedure, and eliminates the need for expensive instrumentations and diagnostic specialists. Given the simple probe design process, and its rapid development, SENSR will be a suitable diagnostic method for emerging infectious diseases.

## Methods

### Materials

SplintR ligase, T7 RNA polymerase, extreme thermostable single-stranded DNA binding protein (ET-SSB), DNase I (RNase-free), and ribonucleotide solution mix were obtained from New England Biolabs (NEB, Ipswich, MA, USA). Recombinant RNase Inhibitor and pMD20 T-vector were obtained from Takara (Shiga, Japan). Malachite green oxalate was purchased from Sigma-Aldrich (St. Louis, MO, USA). Dithiothreitol was acquired from Thermo Fisher Scientific (Waltham, MA, USA). Potassium chloride and 5′-phosphate modified oligonucleotides were obtained from Bioneer, Inc. (Daejeon, Republic of Korea). Full-length probe oligonucleotides were synthesized from Integrated DNA Technologies, Inc. (IDT, Coralville, IA, USA). Oligonucleotides other than these were synthesized from Cosmogenetech, Inc. (Seoul, Republic of Korea).

### Preparation of target RNA

Target RNA was synthesized by an *in vitro* transcription process. To accomplish this, the template DNA containing the target RNA region was amplified by PCR with primer, including the T7 promoter sequence, and cloned into pMD20 T-vector. The PCR amplicon was used as a template for *in vitro* transcription. A transcription reaction mixture containing 1 μg of target DNA, 2 μL 10× T7 RNA polymerase reaction buffer, 1 μL DTT (1 mM), 0.8 μL NTPs (1 mM for each NTP), 0.5 μL Recombinant RNase Inhibitor (20 U), 2 μL T7 RNA polymerase (50 U), and 8.7 μL RNase-free water was incubated at 37 °C for 16 h. The resulting reaction products were treated with 1 μL of DNase I (RNase-free) for 1 h at 37 °C. The transcript was purified using the Riboclear™ (plus!) RNA kit (GeneAll, Seoul, Republic of Korea) and quantified using the absorbance at 260 nm. The purified RNA was used immediately for the downstream reaction or stored at −80 °C. The RNA transcripts were assessed by an Agilent 2100 Bioanalyzer (Agilent Technologies, Santa Clara, CA, USA) using an RNA 6000 nano kit (Agilent Technologies) following the manufacturer’s direction. All primers are listed in Supplementary Table 4.

### Preparation of MSSA and MRSA cell lysates

MRSA (NCCP 15919) and MSSA (NCCP 11488) were obtained from the Korea Centers for Disease Control and Prevention (Osong, Republic of Korea) and cultured for 24 h at 37 °C in 5% sheep blood agar (SBA) (Hanil Komed, Seoul, Republic of Korea). Cells were heat lysed at 95 °C for 2 min.

### Preparation of proxy clinical sample

Human serum was purchased from EMD Millipore Corporation (Temecula, CA, USA). MRSA (NCCP 15919) and MSSA (NCCP 11488) were spiked into human serum. Human serum was diluted at a 1/7 ratio in RNase-free water^9^ and the diluted human serum was heat lysed at 95 °C for 2 min.

### RNA-splinted ssDNA ligation assay

The ligation reaction was performed according to a previously reported method^17^. In summary, 200 nM PP, 220 nM RP, and 220 nM target RNA were added to 4 μL reaction buffer containing 10 mM Tris-HCl (pH 7.4), and 50 mM KCl in RNase-free water. The mixture was heated to 95 °C for 3 min, then slowly cooled to room temperature. This was followed by the addition of 1 μL 10× SplintR buffer and 0.5 μL of SplintR ligase (25 U), and incubation of the mixture at 37 °C for 30 min. The reaction was terminated by heating at 95 °C for 10 min. The ligated product was amplified through PCR reaction with LigChk_F and LigChk_R primers (Supplementary Table 4). The PCR products were assessed by an Agilent 2100 Bioanalyzer using a DNA 1000 kit (Agilent Technologies) according to the manufacturer’s protocol.

### Malachite green and aptamer binding assay

A 1 μM solution of RNA transcript containing malachite green aptamer was mixed with a reaction buffer (50 mM Tris-HCl at pH 7.5, 1 mM ATP, 10 mM NaCl, and 140 mM KCl) to produce 90 μL of solution. The mixture was heated to 95 °C for 10 min and left at room temperature for 20 min. A 5 μL solution of 10 mM MgCl2 was added to the mixture and allowed to stabilize at room temperature for 15 min, followed by addition of 5 μL of 320 μM malachite green solution to produce a total volume of 100 μL. The mixture was incubated at room temperature for 30 min. After incubation, fluorescence intensity was measured using a microplate reader (Hidex, Lemminkäisenkatu, Finland) in 384-well clear flat-bottom black polystyrene microplates (Corning Inc., Corning, NY, USA). For the malachite green aptamer fluorescence, the excitation wavelength was 616 nm with a slit width of 8.5 nm, and the emission wavelength was 665 nm with a slit width of 7.5 nm. Background intensity from the malachite green buffer containing 16 μM malachite green was subtracted from all fluorescence intensities.

### SENSR protocol

The one-step, isothermal reaction master mix consisted of the following components: 2 μL of 1 μM PP, 2.2 μL of 1 μM RP, 5 μL of 320 μM malachite green solution (or 20 μL of 50 μM DFHBI-1T solution), 0.8 μL of ET-SSB, 0.5 μL Recombinant RNase Inhibitor (20 U), 10 μL of SplintR ligase, 5 μL of T7 RNA polymerase (50 U), and 10 μL of 10× SENSR buffer containing 50 mM Tris-HCl (pH 7.4), 10 mM MgCl2, 1 mM of each NTPs, and 10.5 mM NaCl. The reaction master mix was adjusted to 99.22 μL in RNase-free water and 0.78 μL of target RNA was added to produce a total volume of 100 μL. The reaction solution was incubated at 37 °C for 2 hr. After incubation, fluorescence intensity was measured by a Hidex Sense Microplate Reader, as described above. Background intensity from the SENSR buffer containing 16 μM malachite green was subtracted from all malachite green fluorescence intensities. For the Broccoli aptamer fluorescence, the excitation wavelength was 460 nm with a slit width of 20 nm, and the emission wavelength was 520 nm with a slit width of 14 nm. Background intensity from the 10 μM DFHBI-1T solution was subtracted from all DFHBI-1T fluorescence intensities.

For dual detection, we used the following reaction mixture: 2 μL of 1 μM PP1, 2.2 μL of 1 μM RP1, 2 μL of 1 μM PP2, 2.2 μL of 1 μM RP2, 5 μL of 320 μM malachite green solution, 20 μL of 50 μM DFHBI-1T solution, 0.8 μL of ET-SSB, 0.5 μL Recombinant RNase Inhibitor (20 U), 10 μL of SplintR ligase, 5 μL of T7 RNA polymerase (50 U), and 10 μL of 10× SENSR buffer. The reaction master mix was adjusted to 99.22 μL in RNase-free water and added to 0.78 μL of target RNA, producing a total volume 100 μL. The remaining steps are identical to the single target detection.

## Data Availability

The authors declare that all the data supporting the findings of this study are available within the paper and its Supplementary Information. Raw, preprocessed data for Figs. 2-7 and Supplementary Figs. 1-3 are provided at https://doi.org/10.6084/m9.figshare.11941080.

## Acknowledgments

This research was supported by C1 Gas Refinery Program through the National Research Foundation of Korea (NRF) funded by the Ministry of Science and ICT (NRF-2015M3D3A1A01064926). This work was also supported by an NRF grant funded by the Korea government (MSIT) (No. 2018R1C1B3007409). This research was also supported by “Human Resources Program in Energy Technology” of the Korea Institute of Energy Technology Evaluation and Planning (KETEP), which granted financial resource from the Ministry of Trade, Industry & Energy, Republic of Korea (No. 20194030202330).

## Author contributions

J.W.L. conceived the project. C.H.W., S.J., G.S., G.Y.J., and J.W.L. designed the experiment. C.H.W., S.J., and G.S. performed the experiments. C.H.W., S.J., G.S., G.Y.J., and J.W.L. analyzed the results. C.H.W., S.J., G.S., G.Y.J., and J.W.L. wrote the manuscript.

## Competing interests

C.H.W., S.J., G.S., G.Y.J., & J.W.L. have submitted a provisional patent application (No. 10-2019-0046713) relating to the one-step isothermal RNA detection.

